# Effect of short-term heating on bioefficacy of deltamethrin-coated long-lasting insecticidal nets

**DOI:** 10.1101/2021.08.22.21261817

**Authors:** Nakei Bubun, Timothy W. Freeman, Moses Laman, Stephan Karl

## Abstract

We recently reported that long-lasting insecticidal nets (LLINs) distributed in Papua New Guinea (PNG) between 2013 and 2019, exhibited severely diminished efficacy to knock down and kill susceptible Anopheles mosquitoes. This coincided with a rise in malaria observed in PNG since 2015. Here we show that LLIN bioefficacy is increased by heating LLINs prior to WHO cone bioassays.

Unused LLINs with low bioefficacy, delivered to PNG in 2019, were heated to 120°C for 5 minutes. Cone bioassays were performed before and at 1 h, 7 days and 30 days after heating. This led to a significant increase in 24 h mortality (17% to 61%) and 60 min knock down (31% to 72%). The effect was sustained over 30 days.

Bioassays are crucial in quality assurance of LLIN products. Our findings indicate that bioassay results can easily be manipulated. This may have implications for quality assurance procedures used to assess LLINs.

## Introduction

Accounting for more than 50% of the annual global malaria control budget, long lasting insecticidal nets (LLINs) are estimated to have prevented 1.5 billion malaria infections and saved 6.8 million lives between 2000 and 2015 ^1, 2^. Currently, more than 200 million LLINs are distributed to recipient countries per year ^3^. Nevertheless, malaria rates are on the rise again in many countries and, overall, global estimates of morbidity and mortality have stagnated after decades of continuous decline ^2^.

The number of LLIN products available to donors has increased considerably over the last 10 years ^4^. Normally, new candidate LLIN products undergo WHO-supervised assessment in a prequalification (PQ) process. The PQ process involves extensive validation of candidate product bioefficacy, and chemical and physical properties under laboratory and operational conditions ^4, 5^. The most widely used and robust tests to measure LLIN bioefficacy in the WHO prequalification process and in post-marketing surveillance, are standardized WHO cone bioassays in which pyrethroid susceptible mosquito strains are exposed to LLIN samples for 3 minutes ^5^. Adequate bioefficacy is indicated if, after 3 min of exposure to a LLIN that has been washed 20 times, 95% of mosquitoes are knocked down within 60 minutes or 80% of mosquitoes are found dead within 24 h ^5^. Once products have been PQ listed, they are eligible for donor-funded procurement, which drives the vast majority of the LLIN market ^3^. Post marketing, LLIN quality assurance monitoring includes predelivery inspections in which total chemical active ingredient (AI) content, wash retention of the AI, and physical strength of LLINs are tested ^4^.

Despite this quality assurance framework, we have recently shown that PermaNet 2.0®, LLINs with severely diminished bioefficacy were distributed in Papua New Guinea (PNG) between 2013 and 2019, unnoticed by regulatory authorities ^7^. Intriguingly, despite predelivery inspection reports indicating adequate total AI content, the bioefficacy of these LLINs in standardized WHO cone bioassays was low and highly variable with an average 24h mortality of just 39.4% for new nets that had never been used or washed ^7^. A study from Nicaragua, published in early 2021, showed similar deficiencies for the same LLIN product, with strikingly low mortality rates for pyrethroid susceptible mosquitoes (less than 20%) just 6 months after distribution ^8^. These findings stand in stark contrast to the bioefficacy of Permanet 2.0 ® manufactured prior to 2013, which was consistently 100% in cone bioassays, even after extended periods of usage ^9^.

Recently, Yang et al. showed that technical-grade deltamethrin, which is the insecticidal AI in 9 out of 22 currently prequalified LLIN products including PermaNet 2.0® can unfold significantly increased insecticidal potency after short heating for 5 min at 120°C, which is slightly above the melting point of deltamethrin (98°C) ^10^. The authors attributed this effect to a change in deltamethrin structure from crystalline to amorphous.

Inspired by these novel findings and the notion of heat regeneration, sometimes mentioned in documentation associated with LLIN bioefficacy testing, especially after washing ^11^, we performed similar experiments with new and unwashed Permanet 2.0 ® LLINs delivered to PNG in 2019. The LLINs used in this study were the same that exhibited very low bioefficacy in WHO cone bioassays upon arrival in PNG ^7^. The purpose of these experiments was to assess whether heating LLINs in a similar way as described by Yang et al. for the technical grade AI would increase bioefficacy as measured in WHO standard cone bioassays ^10^. Being able to increase bioefficacy of LLIN products using such simple methods may hold opportunities of how to improve the efficacy of these important vector control commodities ^10^. However, the ability to manipulate the outcomes of WHO cone bioassays may also compromise standardized quality assurance processes.

## Materials and Methods

### LLIN bioefficacy tests

WHO cone bioassays were conducted in strict adherence to WHO guidelines ^5^. Briefly, pyrethroid susceptible *Anopheles farauti* colony mosquitoes were exposed to LLIN material for 3 min in WHO standard cones (5 mosquitoes per cone, 20 mosquitoes per LLIN section). A total of n=5 sections (25 × 25 cm) per LLIN were tested (4 sides and roof), i.e., a total of n=100 mosquitoes per net. Overall, n=12 Permanet 2.0® LLINs manufactured in 2019 originating from various batches were tested, resulting in a total of n=60 tested sections (12 nets x 5 sections per net).

### Heating procedure

LLIN sections were heated for 5 min in a laboratory oven set to 120°C. Sections were allowed to cool for at least 1 h before conducting WHO standard cone bioassays.

### Analysis

Outcomes of WHO cone bioassays (24 h mortality and 60 min knock down) were determined by calculating the proportions of exposed mosquitoes either dead (after 24h) or knocked down (after 60 min). Results were averaged for the n=5 sections per net. To determine if there were statistically significant differences before and after heating, Wilcoxon’s matched pairs tests (preceded by a Friedman test), were conducted between baseline and the results obtained on days 0, 7 and 30 after heating.

We observed a substantial, permanent, and statistically significant increase in both 24 h mortality and 60 min knock down after LLINs were heated for 5 min at 120°C. Specifically, while nets prior to heating exhibited a median of 17% (11-84%) 24 h mortality and 31% (13-75%) 60 min knock down, this increased to a median of 61% (34-98%) 24 h mortality and 72% (59-100%) 60 min knock down (Figure 1), respectively, directly after heating, i.e., a 2-3 fold increase. The effect was sustained for 30 days, after which samples exhibited a median of 78% (30-96%) 24 h mortality and 73% (41-96%) 60 min knock down (Figure 1).

**Figure 1.**
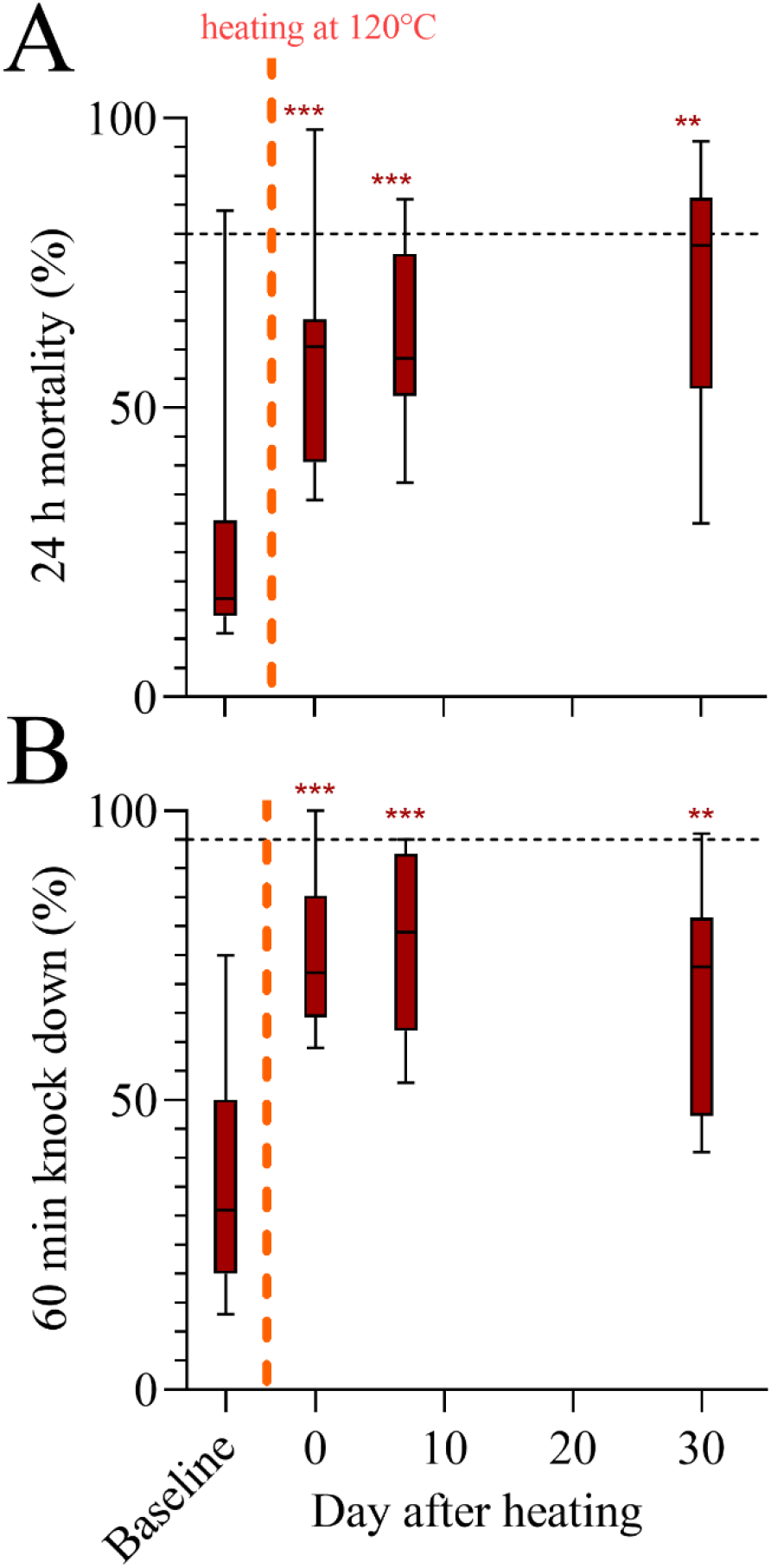
Effect of heating at 120°C on bioefficacy of Permanet 2.0 ® (manufactured in 2019) as observed in WHO standard cone bioassays. Panel A shows 24h mortality before and after heating for 5 minutes. Panel B shows 60 min knock down before and after heating for 5 minutes. Significance is indicated by ** ≤ 0.01; and *** ≤ 0.001 using Wilcoxon matched pairs tests (following Friedman tests) comparing post-heating results back to baseline. Standard Box-and-Whisker plots are presented for the n=12 LLINs per group, showing the median and ranges. The dashed black lines indicate WHO-ratified performance thresholds for LLINs less than 3 years in use or washed ≤ 20 times.

## Discussion and Conclusion

Previous studies have shown that bioassay conditions, including ambient temperature, humidity and the angle of the bioassay cones, can influence mortality and knock down ^12^. The present study shows that the bioefficacy of LLINs as measured using standardized WHO cone bioassays can be substantially altered by heating the net material prior to conducting the assays. This observation has important implications on several levels. Firstly, it is possible that an optimal treatment regimen exists (i.e., time and temperature of heating) that would allow for a complete rescue of bioefficacy in products that are currently observed to exhibit diminished performance. Further research is needed to identify these optimal conditions for various LLIN products. However, it is unknown in how far the heating process affects other important parameters such as wash resistance and physical strength of LLIN products. Thus, further studies would be required to determine whether it is advisable to apply this heating procedure to LLINs that are currently observed to exhibit diminished bioefficacy.

Secondly, our observations imply that bioefficacy can be manipulated by applying simple procedures. Bioefficacy testing is clearly essential in LLIN quality assurance, especially as we have shown that total AI content may not always be correlated with the ability of a product to kill or knock down mosquitoes ^7^. Prequalification procedures and post-market monitoring regulations for LLINs should incorporate this knowledge, ideally by implementing well-validated and standardized methods to measure the surface bioavailable proportion of AI at multiple stages of the product evaluation process, including in recipient countries ^13^.

It is currently unclear what is causing the increased mortality and knockdown in the heat-treated LLINs. Two likely hypotheses are that i) deltamethrin is converted into a more potent form by heating, as recently shown by Yang et al. ^10^. Thus, it could be that the surface bioavailable AI on the LLINs in this study was converted into the more potent form through the application of the heating process. Further studies using electron microscopy-based techniques may be able to elucidate this possibility further.

Alternatively, it is possible that the heating facilitated the diffusion of AI to the LLIN surface thereby increasing surface bioavailability. This has been observed for other LLIN products subsequent to washing, albeit using different temperature and time regimens (e.g, 60°C for 4 h) ^11, 14^. The practical relevance of heat regeneration of LLINs is unclear, as it appears unlikely that end-users in resource-limited settings would widely apply the required procedures subsequent to washing.

In conclusion, here we present thought-provoking evidence that heating LLINs coated with deltamethrin can substantially and permanently increase bioefficacy observed with WHO cone bioassays.

## Data Availability

Data presented in this manuscript will be provided by the corresponding author upon reasonable request.

## Acknowledgments

We would like to thank the staff of the PNG National Malaria Program, Rotarians Against Malaria PNG, and the PNG Institute of Medical Research for their support. SK is supported by an NHMRC Fellowship (GNT 1141441). This study was supported in part by an NHMRC Ideas Grant (GNT 2004390).

## Author Contributions

Conceived Study: SK; Obtained Funding: TWF, ML, SK; Conducted Experiments: NB; Wrote first draft: NB, SK; Supplied samples: TWF; Reviewed manuscript: NB, TWF, ML; SK

